# The effect of walnut consumption on the peripheral blood mononuclear cell transcriptome

**DOI:** 10.1101/2025.10.09.25337286

**Authors:** Alan Kuo, Shan Kurkcu, John W. Birk, Haleh Vaziri, Nuoxi Fan, Daniel W. Rosenberg, Charles Giardina

**Author notes:** Corresponding authors: D.W.R.: Center for Molecular Oncology, School of Medicine, University of Connecticut Health Center, 263 Farmington Avenue, Farmington, CT 06030., C.G.: Department of Molecular and Cell Biology, 91 North Eagleville, Storrs, CT 06269.

## Abstract

Gene expression in peripheral blood mononuclear cells (PBMCs) isolated from subjects before and after a 3-week walnut supplementation was examined for differentially expressed genes by RNA-seq. Significant gene expression changes were observed following walnut consumption with gene enrichment analysis showing significant overlap with genes previously associated with a robust memory B cell response following vaccination. An in-depth clustering analysis of the data revealed that a relatively large subpopulation of the subjects (11/19) showed a stronger and more uniform response to walnut consumption. This group of individuals showed an even more extensive overlap with genes associated with memory B cell production, with a significant overlap also observed for genes activated by an influenza vaccine adjuvant. The genes altered by walnut included numerous kinases and transcription factors, yet a deconvolution analysis did not reveal changes in the major PBMC cell types. Our findings show that a walnut supplementation can change gene expression in PBMCs in a manner consistent with a more responsive adaptive immune response without affecting the cellular composition of the blood.

## Introduction

Nuts are a food category with well-demonstrated health benefits, with walnuts being particularly beneficial. Walnuts provide polyunsaturated fatty acids (PUFAs, ∼47–62% of total fat) with notable levels of α-linolenic acid (ALA), polyphenols (especially ellagitannins like pedunculagin and ellagic acid precursors), dietary fiber, vitamins (e.g., folate, vitamin E in the γ-tocopherol form), minerals (magnesium, potassium), melatonin, and amino acids (particularly L-arginine)**(Ros, 2010)**. Further, walnuts are rich in ellagitannins, a polyphenol metabolized by gut bacteria to bioactive urolithins, which are proving to have several beneficial health effects. While much is known about the general outcomes of walnut consumption, somewhat less is known about the mechanisms underlying these effects **(D’Amico et al.,2021)**. In addition, there may be related health effects that are not yet fully appreciated.

One well documented effect of walnut consumption is a reduced cardiovascular disease (CVD) risk. Retrospective studies of hundreds of thousands of individuals show that walnut consumption significantly lowers CVD risk **(Guasch-Ferre et al.,2017)**. This protective effect is likely due to the ability of walnut to reduce serum LDL cholesterol and blood pressure. The ability of walnut to lower LDL cholesterol is due to their high unsaturated fatty acid content, which reduces cholesterol esterification and increases cholesterol clearance by the liver. The effect of walnut on blood pressure may also be related to their fatty acid content, particularly ALA. Direct administration of ALA to hypertensive rats was found to increase production of vasodilating agents including nitric oxide and bradykinin and decrease systolic blood pressure **(Sekine et al.,2007)**. Walnut is a good source of arginine, the rate limiting substrate for NO synthesis, which is a critical signal for vasodilation and blood pressure regulation.

Walnut consumption has been reported to suppress inflammation in animal models and can decrease circulating inflammatory cytokine levels in humans **(Cofan et al.,2020; Moussa et al.,2025)**. The anti-inflammatory effects of walnut are likely linked in part to changes in prostaglandin metabolism. Many immune and inflammatory cells maintain high levels of arachidonic acid (AA) in their phospholipid membranes, enabling their contribution to prostaglandin synthesis **(Calder, 2008)**. A 4-week walnut supplementation in humans was found to reduce AA in PBMC phospholipids while increasing the level of n-3 fatty acids **(Chiang et al.,2012)**. ALA can also be converted to longer-chain omega-3s EPA and DHA, with known anti-inflammatory effects **(Zhao et al.,2004)**. The impact of walnut on serum lipoprotein particles is also likely to play a role in suppressing inflammatory meditator production. LDL cholesterol incorporates into inflammation-inducing plaque deposits and activates macrophages to increase cytokine production **(Catapano et al.,2017)**, which is reduced by decreased LDL cholesterol following walnut consumption. Other components of the walnut may also potentially suppress inflammation, including vitamin E and the microbial ellagitannin metabolite, urolithin A. Our recent clinical trial has shown that the ability of an individual to produce urolithin A following walnut consumption is associated with the reduction of several inflammatory markers **(Moussa et al.,2025)**. Vitamin E also reduces inflammation, most likely by acting as a lipophilic antioxidant **(Singh et al.,2022)**.

Inflammatory mediators can also impact the adaptive immune response in a variety of ways. A robust immune response to an infection or immunization requires the acute production of inflammatory cytokines. Conversely, cytokines and prostaglandins associated with chronic inflammation can suppress the adaptive immune response. To clarify how walnuts might affect immune cells, we conducted an RNA-seq experiment of PBMCs taken before and after a 3-week walnut supplementation. Here we present an initial analysis of this data.

## Methods

### Subject selection and blood sample collection

Details of the protocol for this study were previously described **(Moussa et al.,2025)**. Briefly, participants were screened to determine eligibility (visit 1). Eligibility requirements included: i) no allergy to walnuts or other tree nuts, ii) no use of antibiotics in the month prior to consenting, and iii) no treatment with steroids or other anti-inflammatory drugs one week prior to the supplementation. After consent, a one-week run-in was performed, when subjects were asked to stop taking dietary supplements and to avoid walnuts and other ellagic acid-containing food sources. Following the run-in, blood was drawn (at visit 2) and participants were instructed to consume 56 grams (2 oz.) of shelled walnuts each day for three weeks. This amount of walnut consumption was based on previous studies where walnut effects were observed **(Baer et al.,2016; Bamberger et al.,2018; Holscher et al.,2018)**. After 3 weeks, blood was drawn again (visit 3). This study was reviewed and approved by the University of Connecticut Health Center Institutional Review Board and Ethics Committee.

### PMBC isolation

Approximately 5 ml of blood was drawn into a K^2^EDTA vacutainer blood tube (Grenier). Whole blood was diluted at a ratio of 5:7 mL with PBS and layered on a Ficoll-Paque (Cytiva) containing centrifuge tube. The gradient was centrifuged at 800x-g for 30 minutes at room temperature and slowed down without deceleration. The resulting buffy coat was collected, transferred to fresh tube, and washed with PBS. The resulting cell pellet was resuspended in cell cryopreservation medium (10% DMSO; 90% FBS) and frozen in an isopropanol bath at approximately -1° C/minute.

### RNA isolation and RNA-seq

Aliquots of frozen PBMCs were rapidly thawed in a 37° C water bath and collected by centrifugation. Cell lysis was performed using a QIAshredder column, and total RNA was purified with the RNeasy Mini Kit (Qiagen) according to the manufacturer’s protocol. RNA concentration was measured using a NanoDrop spectrophotometer (ThermoFisher), and RNA quality was assessed by Agilent TapeStation electrophoresis. All samples used in this study had an RNA integrity number (RIN) ≥ 9. RNA sequencing was performed at the UConn Center for Genomic Innovation using the Illumina NovaSeq platform with 100 bp paired-end reads at a depth of 50M reads per sample.

### RNA-seq data analysis

FASTQ files were trimmed using Trimmomatic, and quality was assessed with FastQC. Reads were aligned to the human reference genome (GRCh38) using HISAT2, and gene quantification was performed with featureCounts. After removing hemoglobin transcripts, differentially expressed genes before and after walnut supplementation were identified using DESeq2. Data analysis and visualization were carried out with PCATools, EnhancedVolcano, and ComplexHeatmap packages. Cell-type deconvolution was performed using the Granulator package.

## Results

### mRNA expression changes before and after walnut consumption

Nineteen volunteers agreed to blood drawings before and after the 3-week walnut supplementation. PBMCs were isolated from the blood, with PBMC aliquots frozen prior to analysis. RNA was prepared from frozen aliquots directly without culturing and subjected to RNA-seq analysis. RNA-seq data was analyzed in duplicate for all but four of the 38 before/after samples (due to RNA yields or sequence quality; Table 1). Sequence data was processed to obtain *FPKM* values, which were then converted to gene counts. The samples were found to have variable levels of red blood cell transcripts, due to RBC contamination, which were removed prior to analysis. The count data for each sample was linked with the subject identification number and whether the sample was taken before or after walnut supplementation. DEseq2 analysis was then performed to identify gene expression changes associated with walnut consumption. Genes with a base mean expression of at least 50 that changed with an adjusted *p*-value of less than 0.05 were identified. These genes are shown in Table 2. An enrichment analysis was then performed on these top 50 genes using the ToppGene tool. No significant enrichment was observed for GO or REACTOME annotations. However, when analyzing data sets generated in previous published studies analyzing PBMCs in healthy individuals, a significant overlap was observed with genes that have been associated with a robust memory B cell response following flu vaccination (Table 3) **(Haralambieva et al.,2016)**.

**Table 1:** Samples For RNA-Seq.

| Table 1: Samples For RNA-Seq |  |  |
| --- | --- | --- |
| Subject | Before Walnut | After Walnut |
| 1002 | Duplicate | Duplicate |
| 1005 | Duplicate | Duplicate |
| 1008 | Duplicate | Duplicate |
| 1009 | Duplicate | Duplicate |
| 1010 | Duplicate | Duplicate |
| 1011 | Duplicate | Duplicate |
| 1012 | Duplicate | Duplicate |
| 1013 | Duplicate | Single |
| 1015 | Duplicate | Duplicate |
| 1016 | Duplicate | Single |
| 1017 | Single | Duplicate |
| 1021 | Duplicate | Single |
| 1035 | Duplicate | Duplicate |
| 1036 | Duplicate | Duplicate |
| 1037 | Duplicate | Duplicate |
| 1038 | Duplicate | Duplicate |
| 1041 | Duplicate | Duplicate |
| 1042 | Duplicate | Duplicate |
| 1043 | Duplicate | Duplicate |

**Table 2:** Gene changes following walnut consumption.

| Table 2: Gene changes following walnut consumption |  |  |  |
| --- | --- | --- | --- |
| Log2 | P-adj | HGNC | Description |
| 2.43 | 0.0001 | FOSB | FosB proto-oncogene, AP-1 transcription factor subunit |
| 1.35 | 0.0014 | EMP1 | epithelial membrane protein 1 |
| 0.99 | 0.0014 | MYADM | myeloid associated differentiation marker |
| 1.60 | 0.0037 | RN7SL5P | RNA, 7SL, Cytoplasmic 5, Pseudogene |
| 1.88 | 0.0039 | RN7SL1 | RNA component of signal recognition particle 7SL1 |
| 1.08 | 0.0056 | RASGEF1B | RasGEF domain family member 1B |
| 1.81 | 0.0059 | RN7SL4P | RNA, 7SL, cytoplasmic 4, pseudogene |
| 0.94 | 0.0065 | PTX3 | pentraxin 3 |
| 1.55 | 0.0065 | ID1 | inhibitor of DNA binding 1 |
| 1.29 | 0.0065 | RN7SL3 | RNA component of signal recognition particle 7SL3 |
| 0.50 | 0.0072 | RAB43-AS | NA |
| 0.59 | 0.0072 | ANXA1 | annexin A1 |
| 1.02 | 0.0087 | TREM1 | triggering receptor expressed on myeloid cells 1 |
| 0.69 | 0.0088 | TUBA1A | tubulin alpha 1a |
| 0.64 | 0.0219 | ARL4A | ADP ribosylation factor like GTPase 4A |
| 0.53 | 0.0219 | PTS | 6-pyruvoyltetrahydropterin synthase |
| 0.82 | 0.0219 | KLF4 | KLF transcription factor 4 |
| -0.35 | 0.0237 | USP11 | ubiquitin specific peptidase 11 |
| 0.26 | 0.0241 | SRF | serum response factor |
| 0.91 | 0.0269 | AHR | aryl hydrocarbon receptor |
| 0.43 | 0.0288 | NUDT15 | nudix hydrolase 15 |
| -0.24 | 0.0297 | DNAJC16 | DnaJ heat shock protein family (Hsp40) member C16 |
| 0.68 | 0.0359 | LDLR | low density lipoprotein receptor |
| 0.22 | 0.0359 | MZF1-AS1 | NA |
| -0.26 | 0.0414 | ATXN2 | ataxin 2 |
| 0.45 | 0.0414 | AKIRIN2 | akirin 2 |
| 0.53 | 0.0481 | ZNF669 | zinc finger protein 669 |

**Table 3:** Gene Enrichment.

| Table 3: Gene Enrichment |  |  |  |  |  |  |
| --- | --- | --- | --- | --- | --- | --- |
| Unclustered subjects |  |  |  |  |  |  |
| GSEA ID | Study description | PubMed | pValue | FDR B&H | FDR B&Y | Bonferroni |
| M41099 | Genes associated with a robust memory B cell response to FLUARIX; day 28 | 27317456 | 3.522E-13 | 1.029E-09 | 8.801E-09 | 1.029E-09 |
| Cluster 1 subjects |  |  |  |  |  |  |
| GSEA ID | Study description | PubMed | pValue | FDR B&H | FDR B&Y | Bonferroni |
| M41099 | Genes associated with a robust memory B cell response to FLUARIX; day 28 | 27317456 | 2.75E-25 | 2.58E-22 | 2.37E-21 | 1.55E-21 |
| M4930 | Genes upregulated following TIV flu vaccine; day 7 | 21743478 | 1.43E-16 | 2.72E-14 | 2.51E-13 | 8.16E-13 |
| M41100 | Genes associated with a robust memory B cell response to FLUARIX; day 3 | 27317456 | 1.34E-14 | 1.57E-12 | 1.44E-11 | 7.52E-11 |
| M40997 | Genes upregulated by AS03 adjuvant (versus PBS) following influenza A/H5N1 vaccine; day 1 | 30566602 | 1.91E-14 | 2.14E-12 | 1.97E-11 | 1.07E-10 |

**Table 3:**
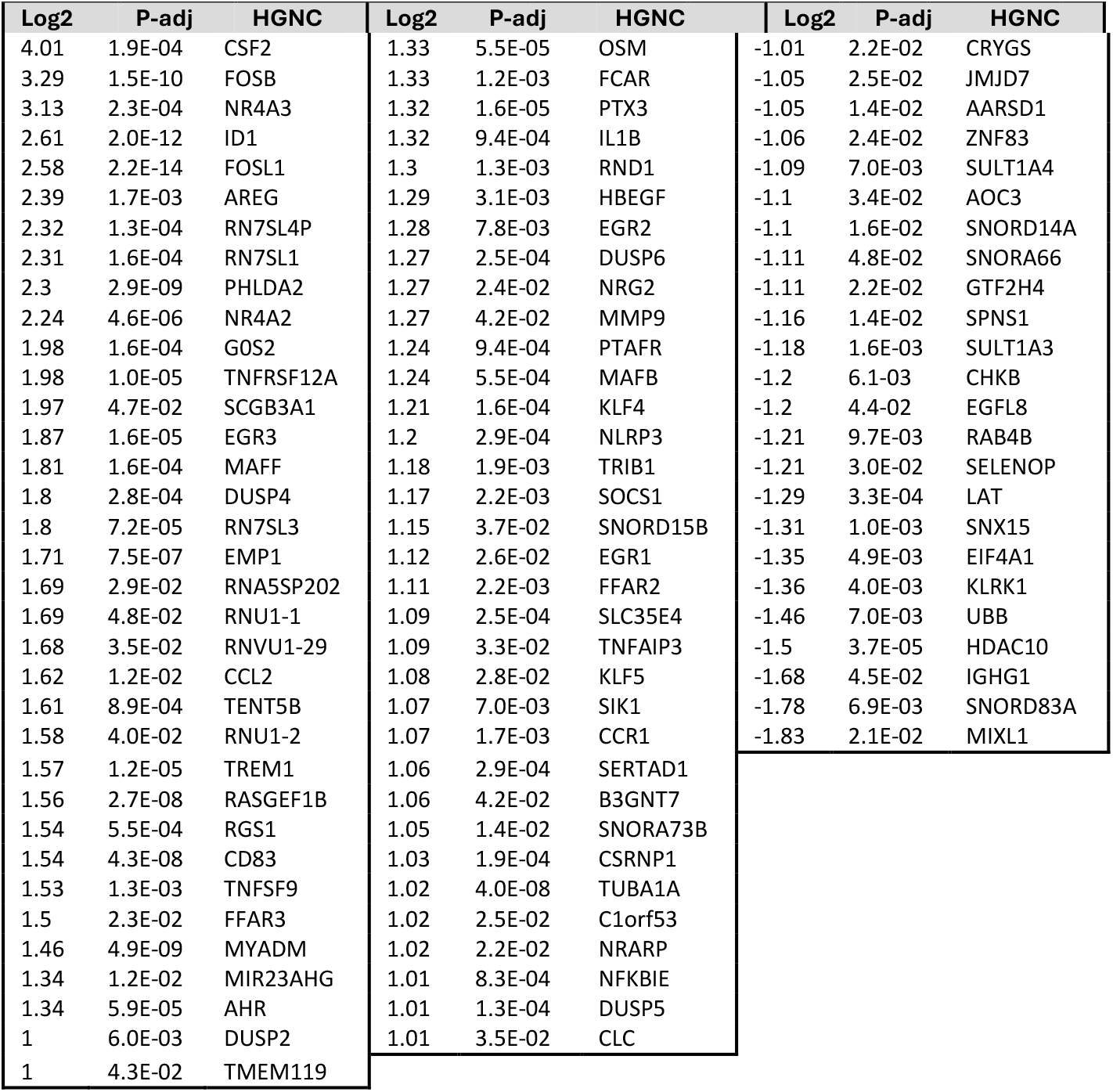
Log2-fold Gene changes in Cluster 1.

### Assessing subject variability to walnut consumption

To assess the interindividual variability of gene expression changes following walnut consumption, a PCA analysis was performed using the PCAtools package based on the genes differentially expressed following walnut consumption (i.e., the genes listed in Table 2). Figure 1 plots the PC1 and PC2 positions of the subjects before (red) and after (blue) walnut consumption. This plot shows that subjects respond similarly to walnut consumption, with a rightward shift in the positive PC1 direction. However, the increased dispersion of the PCA plot following walnut consumption highlights inter-individual variability to this supplementation.

**Figure 1:**
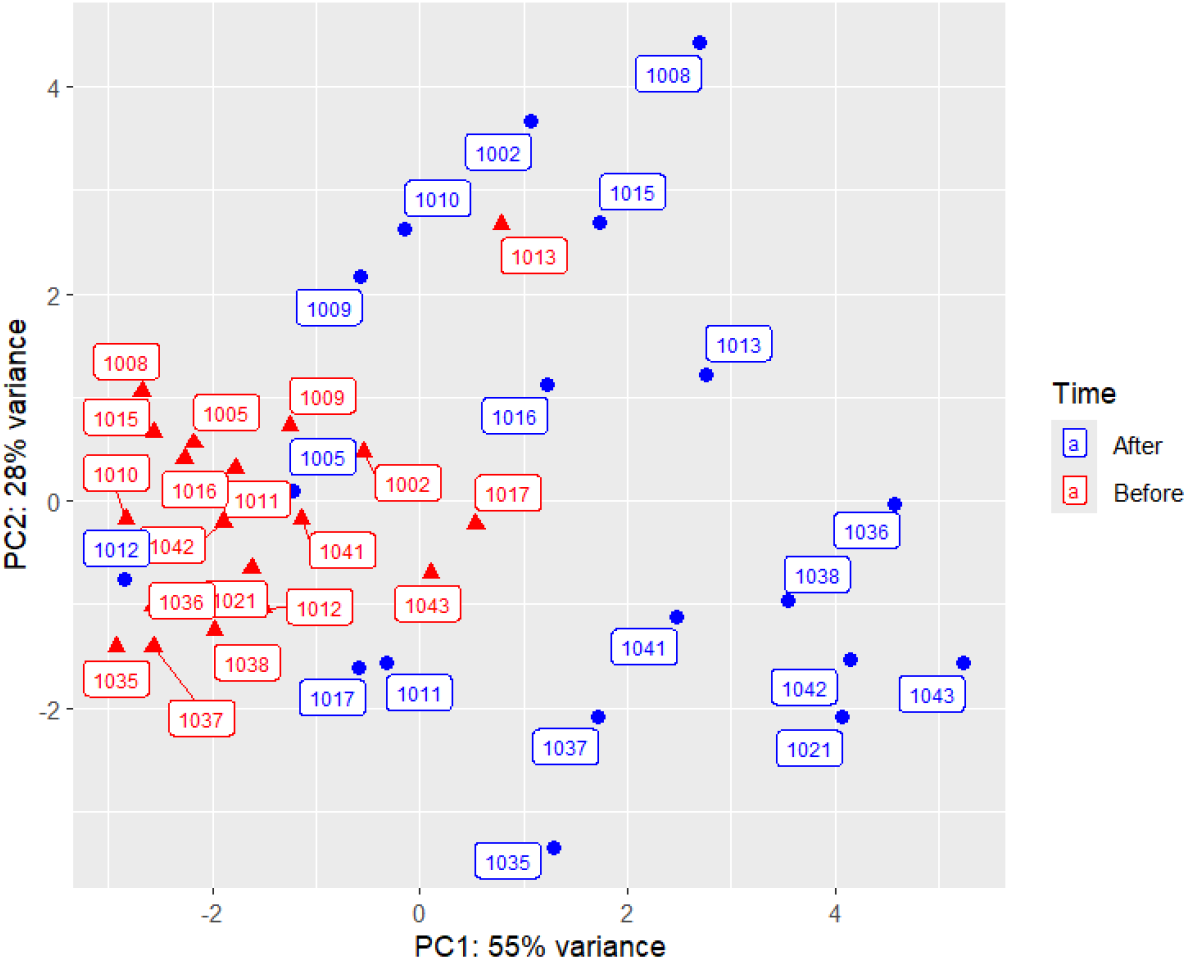
Principal components analysis of subjects before (red) and after (blue) walnut consumption. This analysis was based on the genes in Table 2.

To further assess subject variability, a heatmap was generated using the ComplexHeatmap package, with the subjects in columns and genes listed in Table 2 in rows. Subjects and genes were clustered by ComplexHeatmap’s default clustering function, which uses Euclidian distance to compare the subjects. This analysis sorted individuals into three main clusters (indicated at the top of Figure 2). Clusters 1 and 2 look similar, with the exception that two non-coding RNAs (RN7SL1 and RNSL3) and related pseudogenes (RN7SL5P and RN7SL4P) that are not activated in Cluster 2. Cluster 3 can be broken up into two subclusters, with three individuals showing some similarity to Cluster 1 (1005, 1011 and 1013) and two individuals that don’t respond like the others (1012 and 1017). Overall, these data show some common gene expression changes associated with walnut consumption but also indicate different types of response.

**Figure 2:**
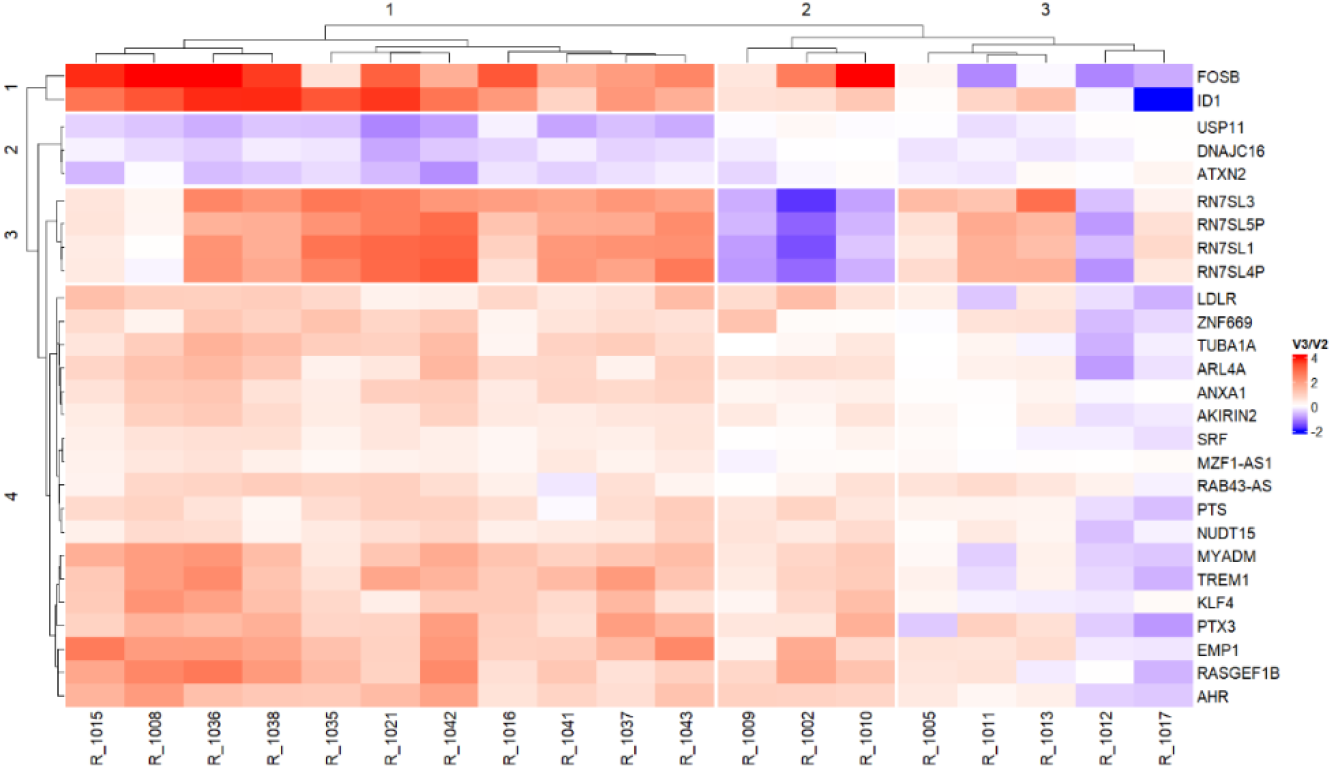
Heat map of gene expression changes following walnut consumption. Red indicates an increase in expression and blue indicates a decrease, as indicated by the scale bar on the right.

Given the variability in the gene expression changes associated with walnut consumption, we analyzed the three clusters shown in Figure 2 independently. When cluster 1 individuals were analyzed, the number of signficantly altered genes more than doubled (Figure 3; Table 3), indicating that this group responded to walnut consumption in a relatively uniform manner. No genes were found to be signficantly changed when clusters 2 and 3 were analyzed separately (Figure 3). As with the overall gene list, enrichment anaysis of the cluster 1 gene list showed similarity with genes associated with a strong memory B cell response to a vaccination (Table 3) **(Haralambieva et al.,2016)**, with this overlap becoming more signficant with the longer cluster 1 gene list. Two other PBMC studies also showed significant overlap with the cluster 1 gene list. One study identified genes more highly expressed following a flu vaccine **(Nakaya et al.,2011)** and the other identified genes stimulated by a vaccine adjuvant that ehances humoral immune responses **(Howard et al.,2019)**. These findings point to an immune-stimualtory effect of walnut consumption.

**Figure 3:**
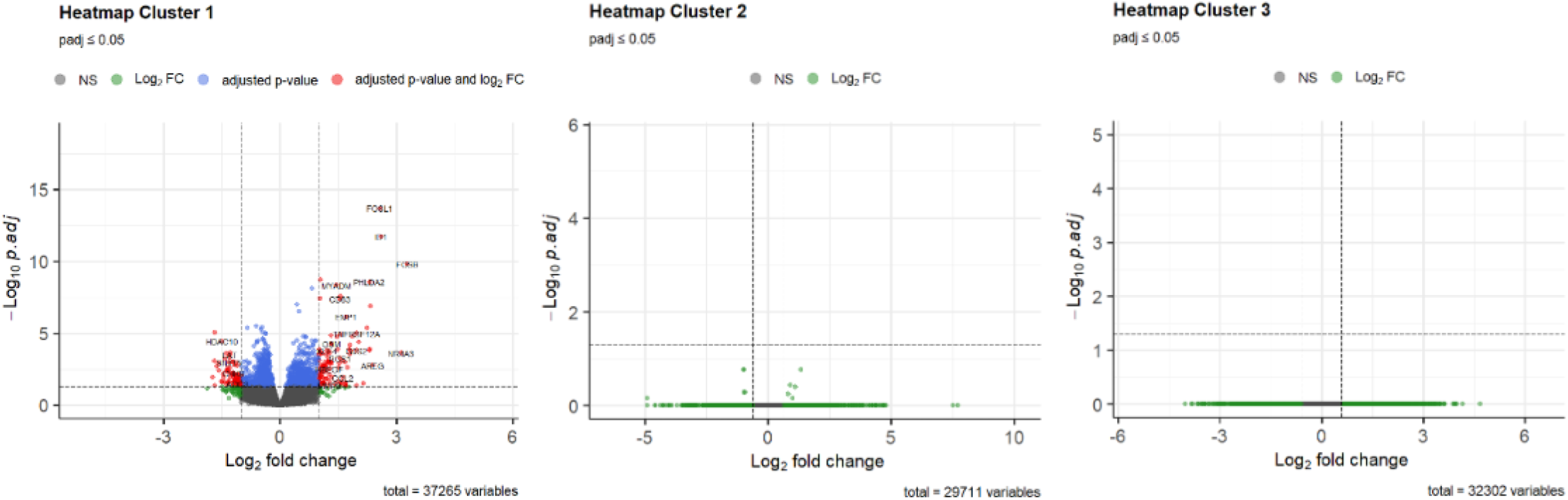
Volcano plot of gene expression changes in clusters 1, 2 and 3 following walnut consumption. Singificantly changed genes are shown in red (p-adj < 0.05).

To further assess subject variability, the “cluster 1” gene set listed in Table 3 was used to generate a heat map using all subjects. This analysis clustered the subjects into three groups. Two groups showed consistent activation of the immune-modulating genes (Groups 1 and 2, Figure 4), with the third group being weakly-responsive (Group 3). However, the two responsive groups 1 and 2 differed in the expression of several non-coding and potentially immunologically-active RNAs: RN7SL1, RN7SL3, RNU1-1, RNU1-2, and RNVU1-29. These RNAs each play a role in protein translation and RNA splicing but can also stimulate an antiviral immune response **(Hoffman et al.,2004; Johnson et al.,2021)**. The increased expression of these non-coding RNAs correlated with a cluster of down-regulated genes (Figure 4). Although the mechanistic connections and physiological consequences of these gene expression differences is not known, this analysis shows variability even in individuals that responded to walnut consumption.

**Figure 4:**
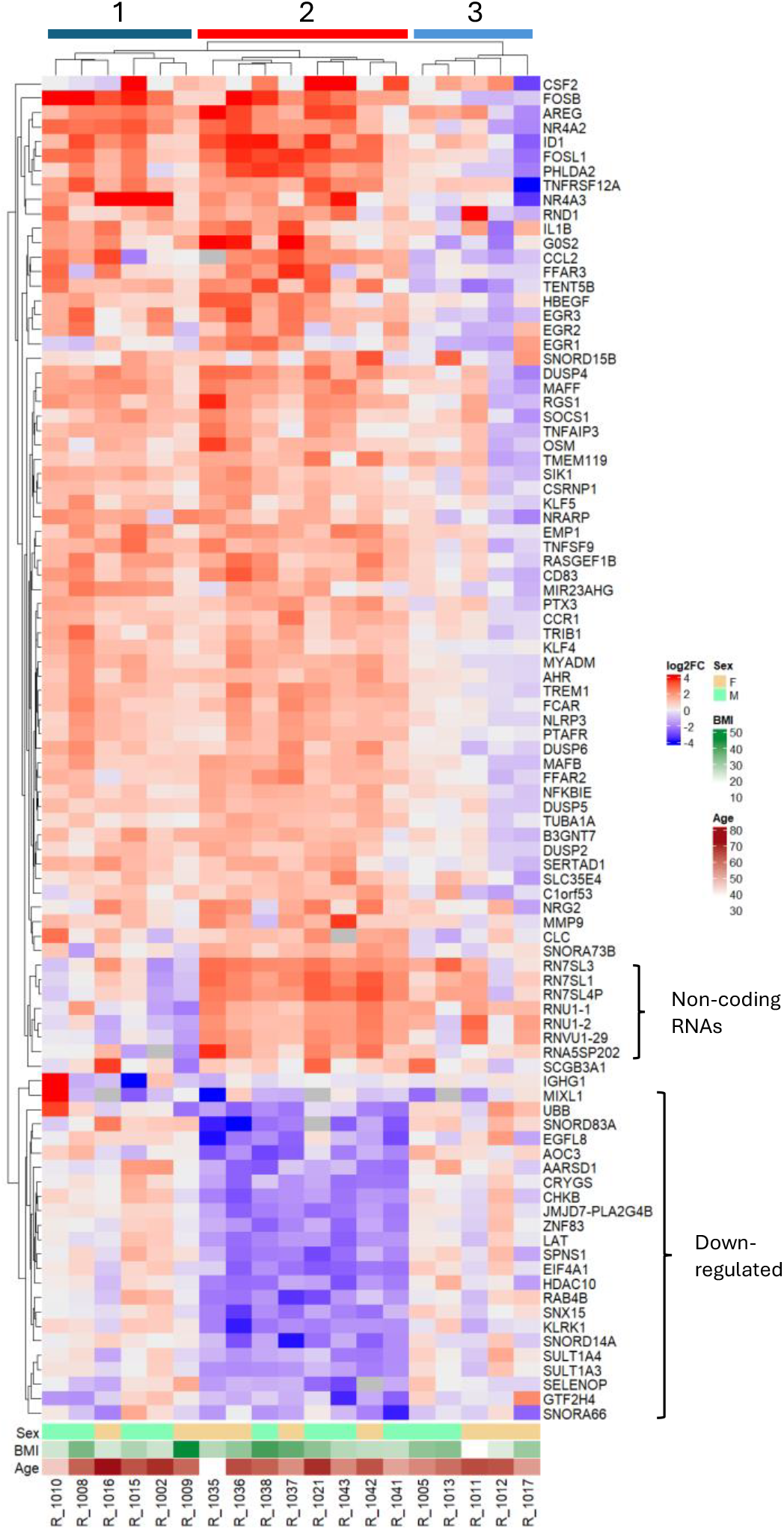
Heat map of gene expression changes following walnut consumption displaying genes with the greatest significance (top 95). Red indicates an increase in expression and blue indicates a decrease, as indicated by the scale bar on the right. Subject sex, BMI and age are shown at the bottom of the graph.

Finally, we determined whether there was an impact of sex, BMI or age on the individual response to walnut. As shown in Figure 4 and 5, no significant effects were observed. There was a trend to higher BMI individuals being in the more responsive clusters 1 and 2, similar to a previous report **(Moussa et al.,2025)**, but this was not statistically signficant (potentially due to the limited number of subjects).

**Figure 5:**
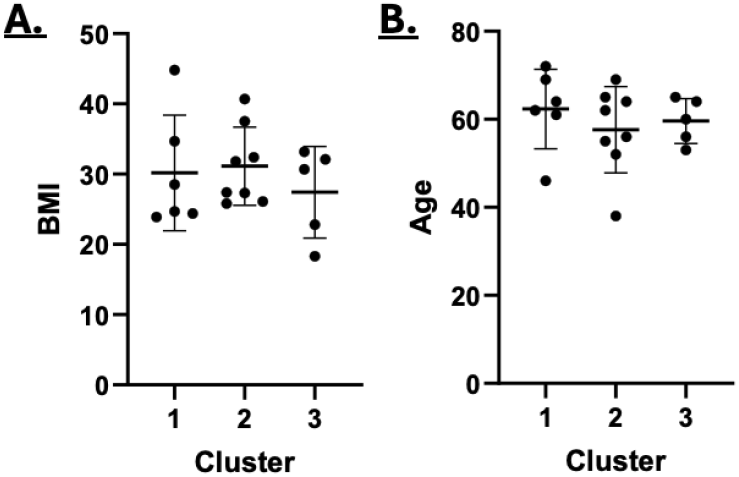
The distribution of subject BMI (A) and age (B) in the three gene expression clusters identified in Figure4. No significant differences were detected.

### Immune cell type deconvolution

To assess the potential effects on the immune cell population, we performed cell-type deconvolution of the bulk RNA-seq data collected before and after walnut supplementation. Deconvolution was conducted using the Granulator R package (version 1.17.0). Because Granulator requires expression values as Transcripts Per Million (TPM), raw RNA-seq counts were first converted to TPM using transcript lengths retrieved from Ensembl, coupled with Granulator’s built-in normalization function. Cell-type proportions were estimated based on the Monaco Immune Data Reference included within the Granulator package. Although the Monaco reference defines 29 immune cell types, our analysis revealed that approximately 95% of PBMCs could be accounted for by just 8 primary cell types (Figure 6). The remaining cell types, such as dendritic cells, neutrophils, and plasmablasts, were excluded due to their negligible representation in the PBMC populations. A Wilcoxon signed-rank test comparing PBMC populations before and after the walnut supplementation showed no statistically significant differences across any of the major immune cell types. These findings indicate that although there are significant changes in gene expression following walnut consumption, the cell population remains stable.

**Figure 6:**
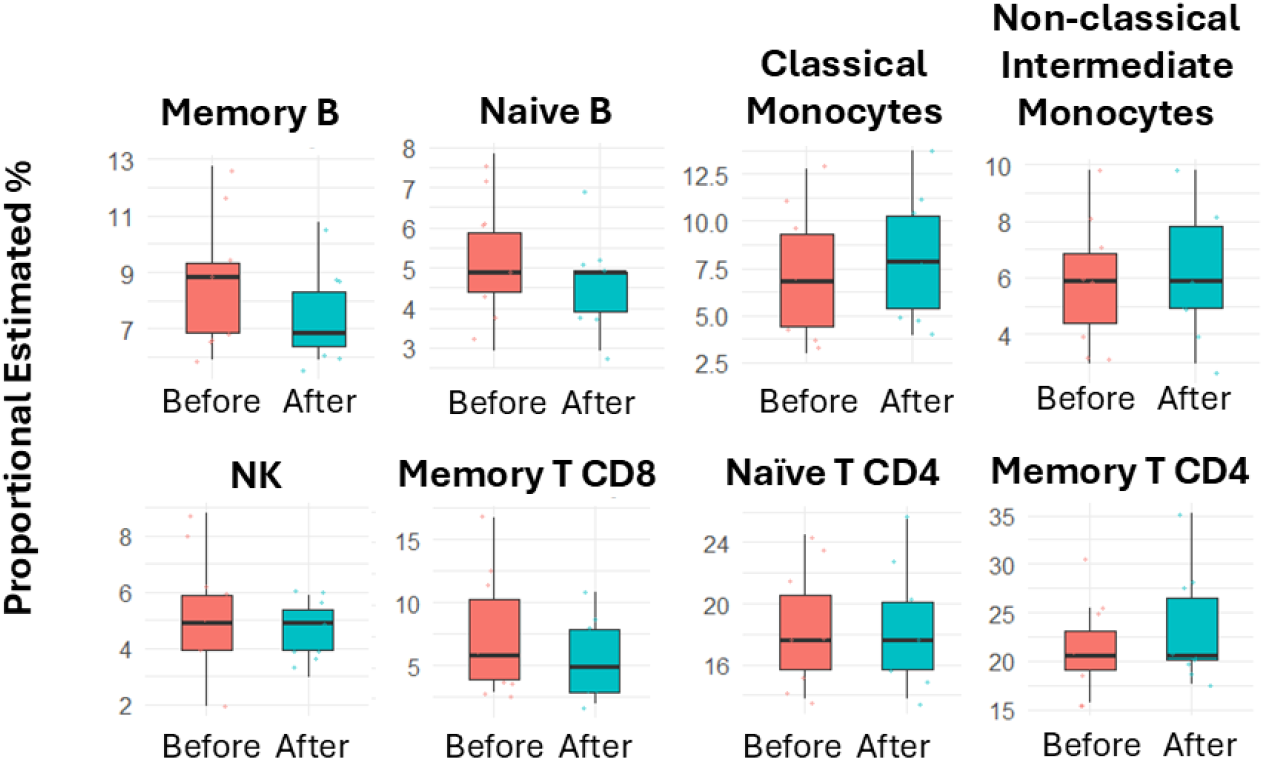
Deconvolution of immune cell changes following walnut consumption. No sigmficant changes were observed.

### Signaling pathway activation

Finally, using the “cluster 1” activated genes listed in Table 3 (derived from the more walnut-responsive individuals), we determined whether there was an enrichment for genes in annotated signaling pathways. Two significant and informative gene annotations were noted. Genes activated by walnut consumption overlapped with the Reactome pathway for “Nuclear events: kinase and transcription factor activation” (Table 4). This enrichment was driven in part by the large number of transcription factors upregulated by walnut, including: FOSB, NR4A3, ID1, FOSL1, NR4A2, MAFF, MAFB, AHR, KLF4, and KLF5. So, although walnut does not impact the PBMC cell composition, it potentially alters the ability of cells to activate genes required for an effective immune response. Another pathway that was significantly enriched was for genes regulated by the transcription factor NFkB following TNF stimulation. This pathway is upregulated in the PBMCs following walnut consumption, even though TNF levels are not increased. An overlapping pathway is presumably responsible for activating these genes. NFkB plays a central role in regulating adaptive and innate immune responses, including the establishment of memory B cells **(Gerondakis & Siebenlist, 2010)**.

**Table 4:** Signaling enrichment.

| Unclustered subjects |  |  |  |  |  |  |
| --- | --- | --- | --- | --- | --- | --- |
| GSEA ID | Category | Type | pValue | FDR B&H | FDR B&Y | Bonferroni |
| M152 | Nuclear events: Kinase transcription factors | Reactome | 4.227E-13 | 3.783E-10 | 2.790E-09 | 3.783E-10 |
| M5890 | TNF signaling via NFkB | Hallmark | 7.710E-39 | 4.290E-35 | 3.947E-34 | 4.290E-35 |

## Discussion

A three-week walnut supplementation by healthy volunteers was associated with changes in the transcriptome of PBMCs. While interindividual response varied between some volunteers, the majority (11 out of 19 subjects) responded similarly. The variation in the response may have been related to other dietary factors, genetic diversity (including microbiome diversity) and/or individual compliance with the study protocol. Genes activated by walnut consumption significantly overlapped with genes previously shown to be associated with a robust memory B cell response following flu vaccination **(Haralambieva et al.,2016)**. These gene expression changes occurred without significant alterations in the cellular composition of the PBMCs. These data suggest that walnut consumption may prime immune cells for a robust response to an infection or immunization in many individuals.

Since our study is limited to a transcriptome analysis, we can only speculate on the physiological impact of the gene expression changes. Gene that regulate immune and inflammatory responses, such as cytokines and chemokines, are tightly regulated even after transcription. For example, we found increased IL1b and CSF2 mRNA expression, without observing higher levels of these cytokines in the blood following walnut consumption. One explanation is that while the cells increase transcription of these mRNAs, they don’t translate and/or release the proteins. This possibility is in line with published data studying IL-1b expression in PBMCs, where distinct transcriptional and translational regulation of IL1b has been demonstrated. In this study mRNA levels were not strictly correlated with protein expression, with IL1b secretion requiring a strong stimulus such as LPS **(Schindler et al.,1990)**. In this regard, we speculate that walnut consumption may help prime the immune response by stimulating transcription of these cytokines and priming cells for an immunogenic stimulus. Interestingly, we did find noncoding RNAs among the walnut activated genes: RN7SL1, RN7SL3, RNU1-1, RNU1-2, and RNVU1-29. RN7SL1 is an interesting finding. This RNA serves a structural function within the signal recognition particle, but it can also be incorporated into extracellular vesicles and transferred to recipient cells, where it activates immune and inflammatory responses via RIG-I binding **(Johnson et al.,2021)**. Likewise, RNU1-1 encodes the U1 snRNA of the U1 snRNP but can also facilitate antiviral innate immunity by promoting TRIM25-mediated RIG-I activation **(Zhang et al.,2024)**. Whether these non-coding RNAs are in fact stimulating components of the innate immune response in PBMCs following walnut consumption is currently unknown.

To gain insight into the potential ramifications of the mRNA expression changes induced by walnut, we evaluated transcriptomic studies of PBMCs from healthy individuals for similar transcriptomic patterns. One gene set that overlapped with the walnut activated mRNAs comes from a study looking for transcriptomic signatures correlating with memory B cell production following vaccination **(Haralambieva et al.,2016)**. In this study participants between 50 and 74 years of age were administered an influenza vaccine. Regression modeling was then performed to identify associations between an influenza-specific memory B cells (by ELISPOT) and PBMC transcriptomic changes at several time points. The walnut-activated genes overlapped most extensively with the last time point taken 28 days following vaccination. Gene enrichment analysis in this study suggested that cholesterol, lipid metabolism, and NF-kB-mediated cytokine signaling, might have caused increased memory B cell production, which is consistent with our finding. Another transcriptomic study with influenza vaccine generated gene sets overlapping the walnut activated genes **(Howard et al.,2019; Nakaya et al.,2011)**. However, in an older study with flu vaccine, some of these same genes were noted to be *reduced*.**(Nakaya et al.,2011)**. The discrepancy between these studies may be related to the characteristics of the study population. Nonetheless, mRNAs that have been associated with immunogenic responses are among those modulated by walnut consumption.

It is interesting that the genes activated by walnut were heavily populated with transcription factors that guide the trajectory of the immune response. These transcription factors include three notable transcription factor pairs: FOSB and FOSL1, NR4A2 and NR4A3 and MAFF and MAFB. FOSB and FOSL1 bind AP1 DNA sequence elements to control the expression of genes that impact multiple phases of the immune response. Recently, it has been found that FOSB, FOSL1 and FOS are selectively expressed in tissue resident memory T cells. These-T cells reside in specific tissue sites to provide localized protection from pathogens **(Smith et al.,2023)**. Whether the increased expression of FOSB and FOSL1 by walnut ultimately impact resident memory T cell formation remains to be determined. The orphan receptors NR4A2 and NR4A3 genes are also highly instrumental in regulating an immune response. NR4A2 regulates immune cell differentiation, favoring the production of Treg cells over Th1 cells, and the production of “anti-inflammatory” M2 over M1 macrophages **(Mahajan et al.,2015; Sekiya et al.,2011)**. NR4A3 plays a key role in steering monocyte differentiation toward dendritic cells rather than macrophages. It also supports the generation of memory T cells and the maintenance of Tregs **(Boulet et al.,2019; Hiwa et al.,2021; Odagiu et al.,2020)**. Among the MAF family of transcription factors, MAFB is known to promote the differentiation of M2 macrophages **(Kim, 2017)**, which are associated with anti-inflammatory responses. In contrast, the immunological role of MAFF remains less well understood. Overall, the observed transcription factor changes suggest a shift toward memory B cell formation and the development of immune-regulatory cell types, such as Tregs and M2 macrophages. Another transcription factor of interest that is activated by walnut exposure is AHR, which can significantly influence immune responses depending on the specific ligands involved **(Gutierrez-Vazquez & Quintana, 2018)**. It is important to note, however, that these findings are based solely on transcriptomic data. Further studies are needed to confirm whether these transcription factors are indeed expressed at the protein level and functionally active in gene regulation.

In summary, we observed a broad spectrum of gene expression changes in human PBMCs following walnut consumption. While these changes suggest potential beneficial effects on the adaptive immune response, further research is needed to clarify the physiological significance of our findings. Understanding the impact of walnut consumption within the complex dietary and genetic backgrounds of humans is challenging. Nonetheless, our findings contribute to the growing body of literature documenting positive effects of walnut consumption on human immune and inflammatory responses.

## Data Availability

All data produced in the present study are available upon reasonable request to the authors

## Acknowledgements

This study was supported by generous funding from the American Institute for Cancer Research /California Walnut Commission to D.W. Rosenberg.

